# The role of socio-economic disparities in the relative success of SARS-CoV-2 variants in New York City in early 2021

**DOI:** 10.1101/2023.09.29.23296367

**Authors:** Tetyana I. Vasylyeva, Jennifer L. Havens, Jade C. Wang, Elizabeth Luoma, Gabriel W. Hassler, Helly Amin, Steve Di Lonardo, Faten Taki, Enoma Omoregie, Scott Hughes, Joel O. Wertheim

## Abstract

**Background:** Socio-economic disparities were associated with disproportionate viral incidence between neighborhoods of New York City (NYC) during the first wave of SARS-CoV-2. We investigated how these disparities affected the co-circulation SARS-CoV-2 variants during the second wave in NYC.

**Methods:** We tested for correlation between the prevalence, in late 2020/early 2021, of Alpha, Iota, Iota with E484K mutation (Iota-E484K), and B.1-like genomes and pre-existing immunity (seropositivity) in NYC neighborhoods. In the context of varying seroprevalence we described socio-economic profiles of neighborhoods and performed migration and lineage persistence analyses using a Bayesian phylogeographical framework.

**Findings:** Seropositivity was greater in areas with high poverty and a larger proportion of Black and Hispanic or Latino residents. Seropositivity was positively correlated with the proportion of Iota-E484K and Iota genomes, and negatively correlated with the proportion of Alpha and B.1-like genomes. The proportion of persisting Alpha lineages declined over time in locations with high seroprevalence, whereas the proportion of persisting Iota-E484K lineages remained the same in high seroprevalence areas.

**Interpretation:** During the second wave, the geographic variation of standing immunity, due to disproportionate disease burden during the first wave of SARS-CoV-2 in NYC, allowed for the immune evasive Iota-E484K variant, but not the more transmissible Alpha variant, to circulate in locations with high pre-existing immunity.

**Funding:** We acknowledge funding from the National Institutes of Health and the Centers for Disease Control and Prevention.

## INTRODUCTION

New York City (NYC), a city of 8.5 million people, was one of the locations most heavily affected by the first wave of SARS-CoV-2 infections in North America and Europe (1, 2). In March through May 2020, >200,000 laboratory-confirmed coronavirus disease 2019 (COVID-19) cases were reported in NYC (2). However, in late March 2020, the percentage of positive PCR tests reached 65% (2), indicating that testing was insufficient to capture the full extent of the outbreak. Most prominently, the epidemiology of SARS-CoV-2 during this first wave reflected socio-economic disparities within NYC, as well as the rest of the US (3), with communities of color and those living in areas with lower average socioeconomic status experiencing the highest burden of SARS-CoV-2 cases, hospitalizations, and deaths (4, 5). The higher proportion of SARS-CoV-2 antibody prevalence in NYC neighborhoods with predominantly Black and Hispanic or Latino residents further highlighted inequalities in access to testing (6). Consequently, after the first wave, NYC neighborhoods were left with drastically different levels of standing immunity, with seropositivity ranging from 11 to 47% in November 2020 at the beginning of the second wave of SARS-CoV-2 infections.

The second wave in NYC, lasting from November 2020 through June 2021, started with an increase in B.1 cases that peaked in early January 2021, followed by a long plateau of high daily number of cases instead of the usual decrease after the peak (7). The plateau was caused by the introduction and co-circulation of a number of variants of concern (VOC) and interest (VOI) that emerged at the time, including the emergence of the B.1.526 lineage, a “local” NYC strain later designated as Iota VOI, characterized by a number of spike mutations that were previously hypothesized to allow SARS-CoV-2 immune-evasive properties (7, 8). Specifically, E484K, D253G, and S477N mutations in the spike protein have been shown to reduce both vaccine-induced and natural immunity. A sister lineage to the Iota variant, currently designated as B.1.637, shared a number of defining mutations (namely L5F, D253G, and T95I), but also included S477N and Q957R. Although prevalent in NYC in early 2021, B.1.637 is not recognized as a VOC or VOI, by the World Health Organization.

Coinciding with the emergence of the Iota variant, the Alpha VOC, which emerged in the United Kingdom in November 2020 (9), was introduced to NYC and started increasing in prevalence. The Alpha variant was characterized by high transmissibility [40-50% higher than the wild-type Wuhan strain (10)] associated with the N501Y mutation that improves its ACE2 receptor binding ability (11). Our previous analysis showed that in January-March 2021, both Alpha and Iota variants grew rapidly in NYC, but a clade of Iota with the E484K mutation (Iota-E484K) outpaced Alpha, B.1.637, and the rest of the Iota lineages in growth (7). At the same time, in the neighboring state of Connecticut and the rest of New York State, regions which were not as heavily impacted by early outbreaks, the Alpha variant was more prevalent compared with Iota (12, 13), as would be expected given the higher transmissibility of Alpha.

Here, we examine how the immune-evasive Iota variant was able to compete and co-circulate with the more transmissible Alpha variant in NYC in early 2021. By focusing on the time that precedes the emergence of other variants and the steep rise in vaccination levels, we examine how differences in seropositivity (primarily from naturally acquired immunity) between different NYC neighborhoods created a niche for successful competition and co-circulation of the two SARS-CoV-2 variants. We examine geographical patterns and socio-demographic characteristics of areas more severely affected during the first SARS-CoV-2 wave in NYC. This analysis sheds light on our understanding of the succession of SARS-CoV-2 variants and the ripple effect of socio-economic disparities on epidemic waves.

## METHODS

### Genetic data

We analyzed 18,332 SARS-CoV-2 genomes sequenced by the NYC Public Health Laboratory (PHL) and the NYC Pandemic Response Laboratory (PRL) from 2 November 2020 through 18 June 2021. Pangolin lineages (v4.0.6) were assigned using UShER (14).

We aligned all Alpha, Iota, B.1.637, and other B.1-like (genomes that were not assigned as any of the other VOC/VOIs or B.1.637) genomes using MAFFT v.7.453 (15). Maximum likelihood trees were inferred using IQTREE2 (16), using the GTR+F+I substitution model, retaining polytomies, and enforcing a minimum branch length of 1x10^−9^ substitutions/site. Within the Iota variant there was a large clade defined by the E484K mutation; we analyzed genomes in this Iota-E484K sub-variant cluster separately from the remaining Iota analyses.

### Seroprevalence level and epidemiological data

Publicly available data collected by the NYC Department of Health and Mental Hygiene and available at https://github.com/nychealth/covid-vaccine-data/ were used to incorporate information on SARS-CoV-2 seropositivity by modified ZIP Code Tabulation Area (MODZCTA), subsequently referred to as ZIP code. We used the seropositivity by ZIP code in NYC as of November 2020 based on the cumulative proportion of positive SARS-CoV-2 antibody tests, available at https://github.com/nychealth/coronavirus-data. This testing method is representative of both natural or vaccine-induced immunity, though vaccination was not broadly available at the start of the second wave in NYC.

Vaccination level by ZIP code was assessed as of May 2021 as the proportion of residents who had received at least one dose of COVID-19 vaccination administered by NYC vaccinating facilities and reported to the Citywide Immunization Registry. Neighborhood poverty level was defined based on the proportion of residents in a ZIP code with household incomes less than the federal poverty level, per the American Community Survey 2014–2018. We defined poverty levels as “Low” (<10% of residents in a ZIP code with household income lower than the federal poverty level), “Medium” (10-19.9%), “High” (20-29.9%), and “Very High” (≥30%). We considered five racial/ethnic groups: Asian or Pacific Islander, Black or African American, Hispanic or Latino, Other (including mixed race, American Indian or Alaska Native, unknown, and other), and White. ZIP code, poverty level, and race/ethnicity data were obtained from the NYC DOHMH population estimates (17).

### Statistical analyses

We performed a linear model regression between the seropositivity in a ZIP code and the proportion of genomes of each studied lineage (e.g., number of Alpha genomes/number of all SARS-CoV-2 genomes reported for that ZIP code) as reported by PHL and PRL. ZIP codes with less than 10 reported genomes were not included in the analysis. Then, we tested the association between seropositivity and poverty at ZIP code level using analysis of variance (ANOVA). We used the Pearson correlation coefficient to test the correlation of the proportion of the population of a certain race/ethnicity in a ZIP code and seropositivity in the ZIP code.

We then calculated the proportion of adjacent ZIP codes (ZIP codes that share a land border) with the similar seropositivity relative to the total number of adjacent ZIP codes and performed a permutation analysis across the NYC ZIP codes, randomly permuting seropositivity 1000 times, to obtain the expected proportion of adjacent ZIP codes with similar seropositivity.

### BEAST analyses

We conducted Bayesian phylogeographic analyses to estimate rates of the variants migration between locations with different standing immunity. We first used BEAST v1.10.4 (18) to reconstruct time-scaled phylogenies for four lineages: Alpha, Iota, Iota-E484K, and B.1.637. We used the GTR+G4 nucleotide substitution model, assuming a strict molecular clock model (fixed to the value of 8×10^−4^ substitutions/site/year), and a Skygrid coalescent tree prior with 20 dimensions to reconstruct past population dynamics (19). We ran Markov Chain Monte Carlo (MCMC) for 10^8^ generations and investigated the chain convergence using Tracer v1.7.1 (20).

For phylogeographic analysis, we used LogCombiner (18) to resample the obtained tree distributions at a lower frequency. The resulting tree distributions for each lineage included 2000 trees; these tree distributions were used for all consequent phylogeographic analyses. We assigned geographic trait value to sequences in the phylogeographic analysis to locations based on the seropositivity reported in the area separately for each of the variants. We ran MCMC for 10^8^ generations to estimate pairwise migration rates between different locations in this analysis.

To assess whether certain variants were preferentially migrating from one location to another based on the seropositivity, we calculated the pairwise difference in seropositivity and estimated whether it was correlated with the log-transformed viral lineage migration rate between the locations. We then re-ran the analyses separately including seropositivity and vaccination level in each location as of May 2021 as a covariate for the migration rates directly in the phylogeographic analysis. We ran these analyses separately for each variant using a previously described approach (21).

### Persistence analysis

We used PersistenceSummarizer, as implemented in BEAST (22), to examine the effect of seropositivity on the persistence of specific variants in a location. PersistenceSummarizer identifies lineages that have been in a location from the beginning to the end of an observation period. For this analysis, we partitioned the observation period into biweekly intervals going backward in time from the most recent genome in each of the datasets.

### Ethical considerations

The institutional review board of UC San Diego granted ethical approval of this work as human subjects exempt.

## RESULTS

### Geographical patterns in seropositivity

To test whether there was geographical structure in the levels of preexisting immunity to SARS-CoV-2 across NYC as of November 2020, we analyzed whether areas with similar seroprevalence levels were more likely to be geographically adjacent to each other. First, we assigned every ZIP code across NYC into one of three quantiles (“High”, “Medium”, and “Low”) based on their reported seroprevalence levels (Fig. 1). We found the proportion of adjacent ZIP codes (i.e., ZIP codes sharing a land border) with the same seroprevalence level was higher than expected and had a strong geographical pattern. Specifically, 20.6% of ZIP codes adjacent to ZIP codes with “High” seroprevalence level also had “High” seroprevalence, the expected proportion was 7.2% (95% range 4.2–10.5%). For “Medium” seroprevalence level ZIP codes, the observed proportion of adjacent ZIP codes in the same category was 45.9%, while the proportion expected by chance was 29.1% (95% range 24.1–34.1%). For “Low” seroprevalence level ZIP codes, the observed proportion of adjacent ZIP codes with “Low” seroprevalence level was 50.7%, while the expected was 26% (95% range 20.8–30.7%). For each seroprevalence level the average proportion of adjacent ZIP codes that match the seroprevalence level was higher than expected, indicating that seroprevalence level in NYC has geographic structure.

**Figure 1.**
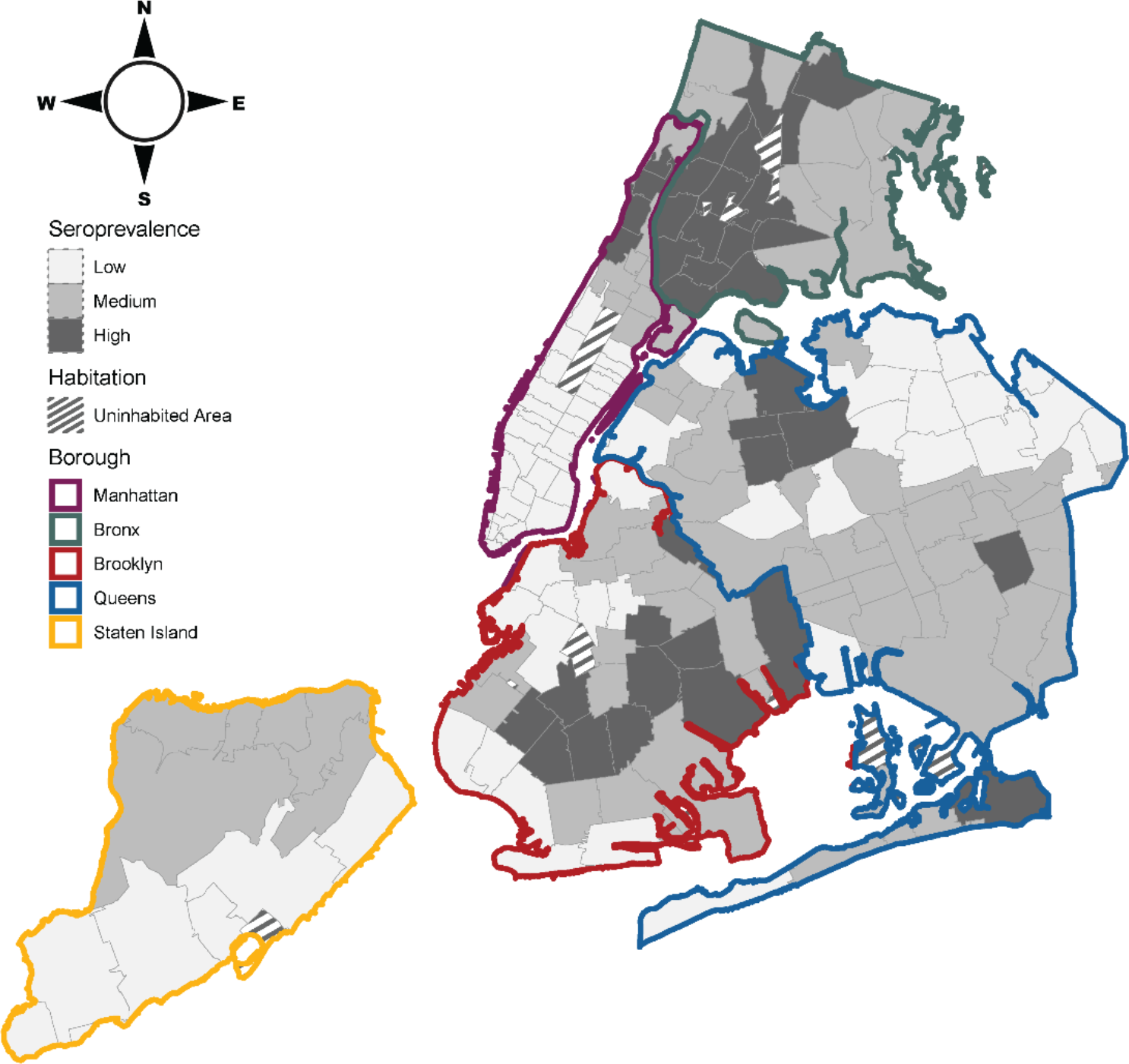
Seroprevalence of SARS-CoV-2 antibodies across NYC in November 2020. Color borders indicate boroughs. Gray borders denote ZIP codes and shading indicates seroprevalence.

### Correlation of variant prevalence and seropositivity

We estimated whether prevalence of SARS-CoV-2 lineages during the second wave was correlated with estimated seropositivity at the start of the second wave, by ZIP code. We focused on Alpha, Iota, Iota-E484K, and B.1.637 lineages and compared them to the co-circulating B.1-like viruses. In locations with higher pre-existing seropositivity, we observed a higher proportion of Iota and Iota-E484K genomes, but not other variants or B.1-like lineages (Fig. 2A). Specifically, we found a positive correlation between the higher pre-existing seropositivity and the proportion of Iota-E484K genomes (R^2^=0.104, *p*<0.001) and Iota genomes (R^2^=0.037, *p*=0.014). In contrast, we found a negative correlation with pre-existing seropositivity and the proportion of Alpha (R^2^=0.026, *p*=0.040) and B.1-like genomes (R^2^=0.082, *p*<0.001); no correlation was found for the B.1.637 lineages (R^2^=0.005, *p*=0.376) (Fig. 2B). Although the correlations of seropositivity and some variants are significant, seropositivity does not explain most of the variance in lineage proportion. We note that ZIP codes with low numbers of sampled genomes will have high variance in estimated lineage proportion and high stochasticity in population dynamics which are other major factors that impact estimated lineage proportion in a ZIP code, along with relative susceptibility of a population to different variants due to standing immunity levels.

**Figure 2.**
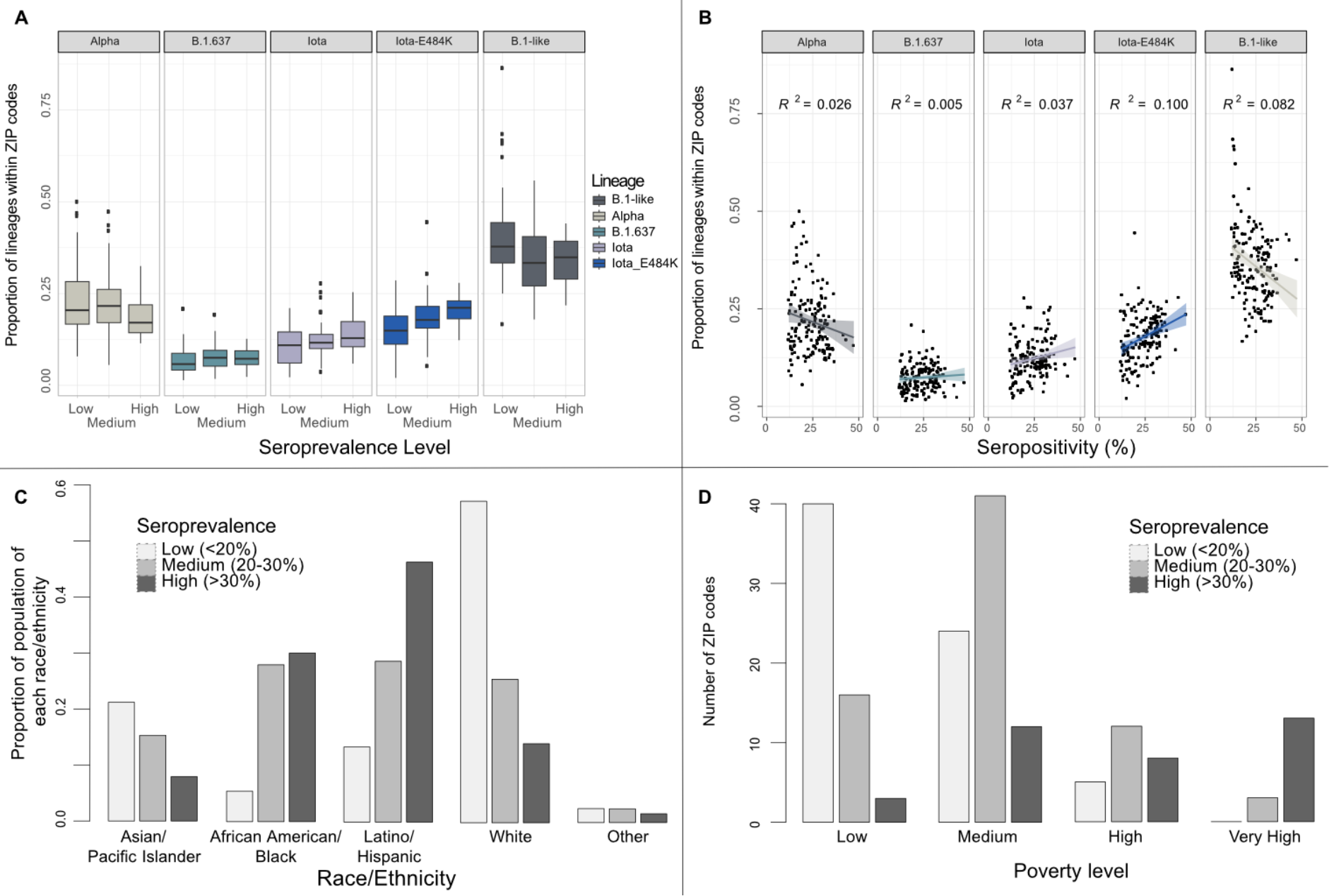
SARS-CoV-2 variants and demographics across ZIP codes in NYC. Correlation analysis between (A) seroprevalence level and proportion of each lineage, (B) seropositivity and proportion of each lineage; (C) proportion of population of a certain race/ethnicity by seroprevalence level by ZIP code, and (D) number of ZIP codes of different seroprevalence level by poverty level.

### Correlation between seropositivity and socio-demographic data

We then investigated the relationship between ZIP code seropositivity and socio-demographic composition (Fig. 2C and Suppl. Table 1). We found a negative correlation between the seropositivity and the proportion of residents who identified as White, Asian or Pacific Islander, or Other races (Pearson correlation coefficients -0.73 (*p*<0.001), -0.31 (*p*<0.001), and -0.23 (*p*=0.002), respectively). Conversely, seropositivity was positively correlated with the proportion of Black or African American and Hispanic or Latino residents (Pearson correlation coefficients 0.49 (*p*<0.001), and 0.63 (*p*<0.001), respectively). Similarly, we found that areas with “High” and “Very High” poverty levels were more likely to have high seropositivity (ANOVA *p*<0.0001; Suppl. Table 1, Fig. 2D).

### Phylogeographic analysis findings

We performed phylogeographic analysis to infer viral lineage migration rates between regions of seroprevalence levels. To make our approach computationally tractable, we grouped all ZIP codes with the same seroprevalence level (“High”, “Medium”, or “Low”; Fig 1) in the same borough into one category and manually reviewed (Fig.3). Since there was only one “Low” seroprevalence ZIP code in the Bronx, it was included in the Bronx “Medium” seroprevalence group. Staten Island did not have any ZIP codes with “High” seroprevalence level, resulting in a total of 13 total locations for phylogeographic inference.

**Figure 3.**
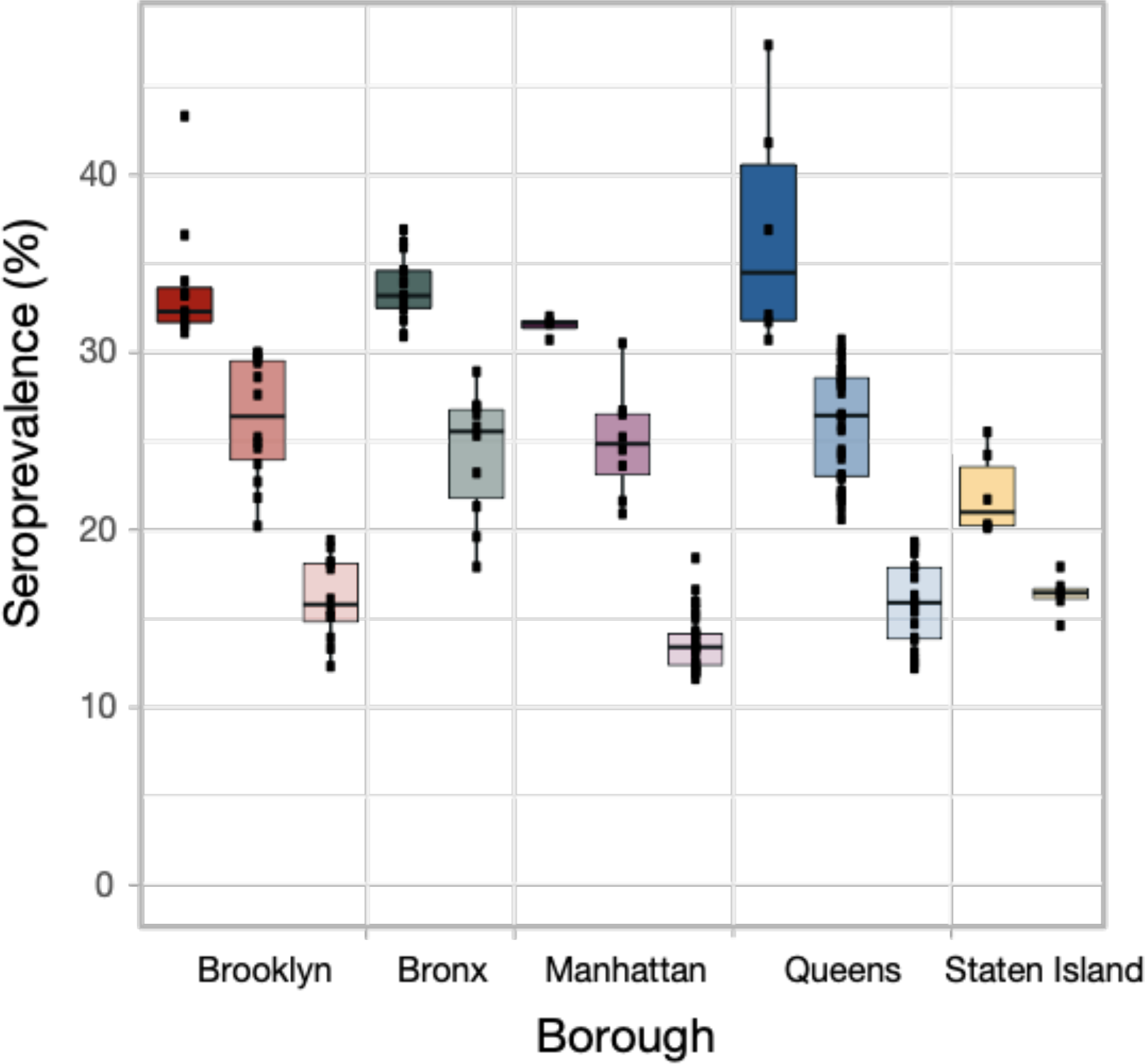
Seroprevalence quantiles across NYC boroughs. Seroprevalence level for ZIP codes (black square); categorized by location groups (high, medium, and low seroprevalence within each borough). Location group summarized by boxplot where shading indicates seroprevalence level, and color is associated with borough.

We found no significant relationship between the log-transformed between-location viral migration rates and the pairwise seropositivity difference for any of the studied lineages: correlation coefficients 0.04 (*p*=0.65), 0.02 (*p*=0.78), 0.17 (*p*=0.84), and -0.06 (*p*=0.48) for Alpha, B.1.637, Iota, and Iota-E484K, respectively. There is no significant pattern of migration between locations that is explained by the difference in seroprevalence of those locations.

Inclusion of seropositivity as a generalized linear model covariate directly in the phylogeographic model also showed no statistically significant association with the migration rates (Suppl. Table 2). The same result was observed when all locations with the same seropositivity level were merged together irrespective of the borough. Additionally, to test whether different levels of vaccination throughout NYC could have affected SARS-CoV-2 migration rates between the different locations as estimated in the phylogeographic analysis described above, we included vaccination level (as of May 2021) as a covariate in our analyses, but similarly, no association was found.

### Analysis of persisting lineages

We further ran lineage persistence analyses that allow inference, whether a variant was likely to persist in a location (i.e., if a variant observed in a location is still present in that same location at the end of the two-week observation period) or if it was more likely to migrate to another location or go extinct (i.e., not observed after two weeks). We ran the persistence analysis separately for each of the variants to describe patterns of their circulation within locations of “High”, “Medium”, and “Low” seroprevalence level. We found that for the Alpha variant, the proportion of lineages persisting in “Low” seroprevalence level locations grew over time, from 19% in mid-January to 81% in early June. Concurrently, the proportion of Alpha lineages was gradually declining in “High” seroprevalence level locations, from 16% to 2% in the same observation period (Fig. 4A). This finding indicates that Alpha was able to consistently persist for longer than 2 weeks in locations with “Low” seroprevalence level, but repeatedly and rapidly went extinct in “High” seroprevalence locations. Given that seropositivity was not a significant predictor for migration rates between locations with different seroprevalence levels, it is likely that Alpha was going extinct in areas with “High” seroprevalence.

**Figure 4.**
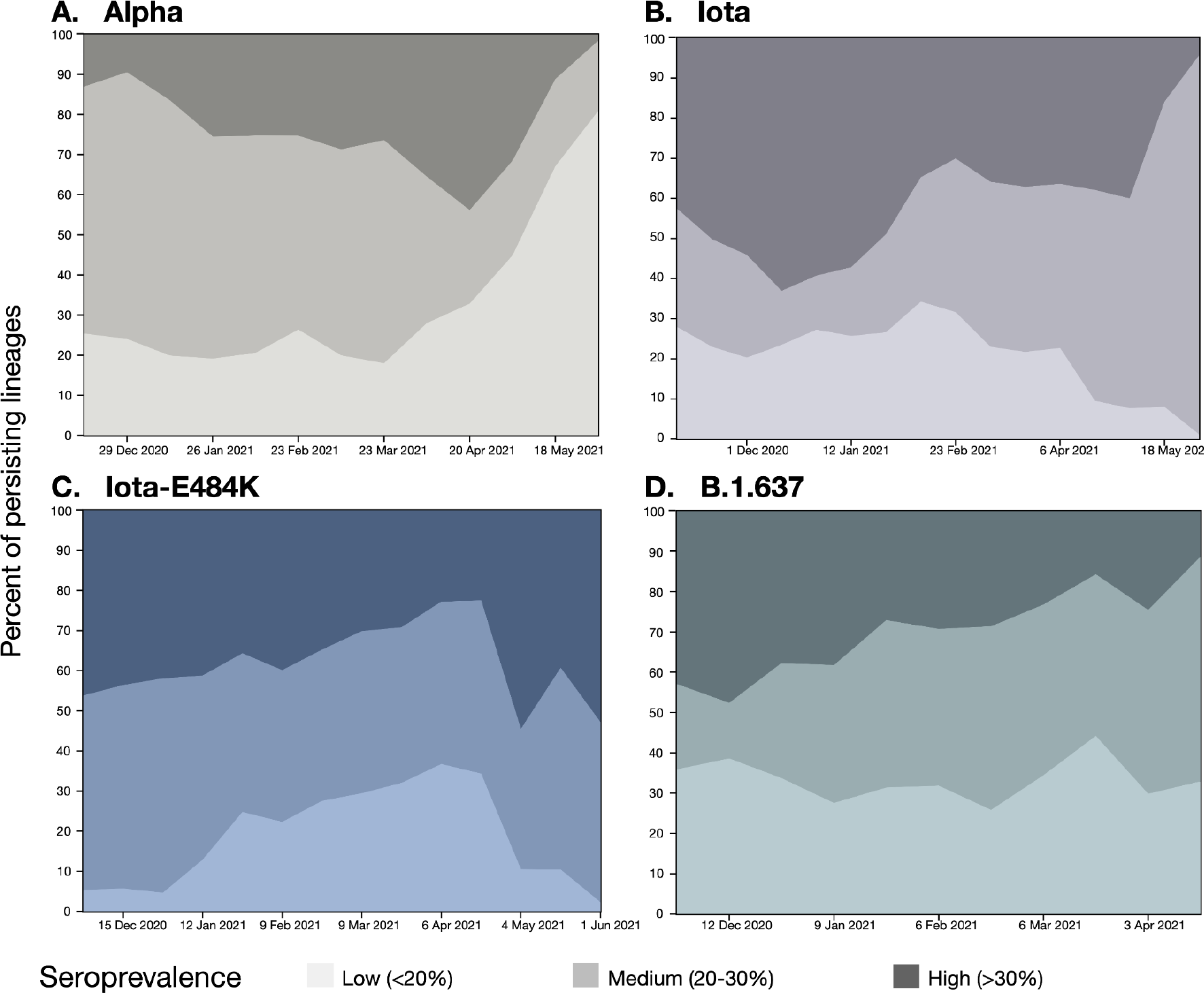
Persistence of variant lineages over time in NYC stratified by seroprevalence level. Percent of persisting lineages indicates the proportion of lineages present in a location at the observation time that were already present in the same location 2 weeks prior. Shading indicates seroprevalence level.

In contrast, the proportion of persisting Iota-E484K lineages remained relatively stable in all locations in the corresponding time period. Between mid-January and mid-May, the proportion of Iota-E484K lineages that persisted in “High” seroprevalence level locations for at least 2 weeks changed from 41% to 39%, and in “Low” seroprevalence level locations from 13% to 10% (Fig. 4B). Importantly, Iota-E484K was not able to establish itself at high prevalence in “Low” seroprevalence level locations, likely in part because they were being outcompeted by infections with the more transmissible Alpha variant.

Other Iota lineages circulated mostly in locations with “Medium” seroprevalence level: the proportion of Iota lineages persisting in both “High” and “Low” seroprevalence level locations was gradually decreasing, while the proportions of Iota lineages persisting in the “Medium” seroprevalence level locations grew from 26% to 96% between mid-January and early June (Fig. 4C). Between mid-January and mid-April, the proportion of B.1.637 persisting lineages remained stable in the “Low” seroprevalence level locations (28% and 33%), grew in the “Medium” seroprevalence level locations from 34% to 55%, and declined in the “High” seroprevalence level locations from 38% to 11% (Fig. 4D).

## DISCUSSION

Infectious disease thrives on disparity and inequality (3). Here we demonstrate how socio-economic disparities in New York City created unequal predispositions for successful spread of emerging lineages during the second wave of SARS-CoV-2. We show that those areas that had high seropositivity after the first wave of SARS-CoV-2 and were predominantly high poverty areas with majority Black and Hispanic or Latino residents, were less likely to sustain the highly transmissible Alpha variant. In contrast, these locales were more likely to sustain the Iota-E484K variant; areas with high levels of preexisting seropositivity provided a successful niche for the emergence and spread of an immune evasive variant (in comparison to a highly transmissible but less immune evasive variant) likely due to re-infection of individuals who had acquired SARS-CoV-2 during the first wave.

Our findings confirm that areas with high seropositivity were more likely to have SARS-CoV-2 infections of Iota-E484K than Alpha, as hypothesized earlier (13); and that once Iota-E484K reached those areas of high seropositivity, it was able to persist there, while Alpha was not. Interestingly, we did not find a correlation between virus migration rates and difference in seropositivity between locations. This finding suggests that when the Alpha variant reached “High” seroprevalence areas, it likely went extinct there, unable to re-infect individuals, but did not successfully migrate to other areas. In contrast, in “Low” seroprevalence locations, Alpha prevalence was able to grow and thus maintain its dominant position in the city.

In May 2021, after the second wave, the proportion of partially vaccinated people in NYC ranged from 22-82% and only 5-17% were fully vaccinated. Vaccination level at that point in time was not a main contributing factor to relative variant prevalence during the second wave that took place in November 2020 through June 2021, as evidenced by our phylogeographic analysis. Less than 1% of people were vaccinated at the start of the second wave.

In NYC, the introduction of the Alpha variant around the time of Iota emergence (7) likely prevented Iota from establishing at a level that would make it difficult for Alpha to compete. Unlike previous analyses that focused on competition at a country level (23, 24), we were able to decipher competition dynamics between variants at the neighborhood level owing to the granularity of seropositivity data and SARS-CoV-2 genome data. Given the higher transmissibility of the Alpha variant, it might have completely eliminated Iota from NYC if not for the pockets of higher seropositivity left in the wake of the first wave of SARS-CoV-2. For example, when the Gamma variant, another lineage with immune-evasive properties, reached NYC in January 2021 (25), it was not able to successfully establish in the city and only circulated at low frequencies, likely because its niche in people with pre-existing immunity was taken by Iota-E484K that had been circulating in NYC for several months.

The Gamma variant dominated the Brazilian SARS-CoV-2 epidemic in late 2020/early 2021 due to the high levels of pre-existing immunity in Brazilian population (26, 27), but in mid-2021 it was quickly replaced by Delta due to the Delta variant’s higher transmissibility (28). At the same time, the Gamma variant was not able to successfully establish itself in the neighboring Colombia or Peru (23, 24). This failure is likely due to a founder effect as competing variants, Mu in Colombia and Lambda in Peru, which also have immune-evasive properties, already dominated the Colombian and Peruvian epidemic by the time Gamma reached the country. Thus, the time of VOCs emergence and introductions, and mechanisms for increased fitness, is crucial for their success in a location in the presence of other variants.

Our study was able to elucidate how various factors, including geographic variability in seropositivity, a symptom of inequities associated with socio-demographic factors in 2020, affected the competition dynamic between two co-existing viral lineages. Such analyses would not be possible in the post-Omicron era when multiple consecutive waves are dominated by variants with similar properties and where the majority of the population have a mixture of acquired natural and/or vaccine-induced immunity (29). By focusing on the time preceding the emergence of the Delta and Omicron variants, we show that the consequences of socio-economic disparities in an outbreak can have a ripple effect that can last into subsequent outbreaks, providing an important lesson for future epidemic preparedness and mitigation efforts.

## Data Availability

All data produced in the present study are available upon reasonable request to the authors.

## Acknowledgements

NYC DOHMH: Public Health Laboratory: Virology Section, WGS Unit; Bureau of Communicable Diseases for COVID-19 seroprevalence data; Division Management Systems Coordination for data integration; Bureau of Immunization for COVID-19 vaccine data; NYC DOHMH Bureau of Epidemiology Services for NYC MODZCTA data and shapes; Pandemic Response Laboratory.

This work was supported (in part) by the Epidemiology and Laboratory Capacity (ELC) for Infectious Diseases Cooperative Agreement (Grants: ELC DETECT and ELC DETECT Expansion) funded by the Centers for Disease Control and Prevention (CDC). Its contents are solely the responsibility of the authors and do not necessarily represent the official views of CDC or the Department of Health and Human Services.

## References

1. Gonzalez-Reiche AS, Hernandez MM, Sullivan MJ, Ciferri B, Alshammary H, Obla A, et al. Introductions and early spread of SARS-CoV-2 in the New York City area. Science. 2020;369(6501):297–301.

2. COVID-19 Outbreak — New York City, February 29–June 1, 2020 Weekly / November 20, 2020. 2020;69(46):1725–9.

3. Mude W, Oguoma VM, Nyanhanda T, Mwanri L, Njue C. Racial disparities in COVID-19 pandemic cases, hospitalisations, and deaths: A systematic review and meta-analysis. J Glob Health. 2021;11:05015.

4. CDC. Rapid Emergence and Epidemiologic Characteristics of the SARS-CoV-2 B.1.526 Variant — New York City, New York, January 1–April 5, 2021. Weekly. 2021;70(19):712–6.

5. CDC. Notes from the Field: Epidemiologic Characteristics of SARS-CoV-2 Recombinant Variant XBB.1.5 — New York City, November 1, 2022–January 4, 2023. Weekly. 2023;72(8):212–4.

6. Parrott JC, Maleki AN, Vassor VE, Osahan S, Hsin Y, Sanderson M, et al. Prevalence of SARS-CoV-2 Antibodies in New York City Adults, June-October 2020: A Population-Based Survey. J Infect Dis. 2021;224(2):188–95.

7. West AP, Jr., Wertheim JO, Wang JC, Vasylyeva TI, Havens JL, Chowdhury MA, et al. Detection and characterization of the SARS-CoV-2 lineage B.1.526 in New York. Nat Commun. 2021;12(1):4886.

8. Thompson CN, Hughes S, Ngai S, Baumgartner J, Wang JC, McGibbon E, et al. Rapid Emergence and Epidemiologic Characteristics of the SARS-CoV-2 B.1.526 Variant - New York City, New York, January 1-April 5, 2021. MMWR Morb Mortal Wkly Rep. 2021;70(19):712–6.

9. Walker AS, Vihta KD, Gethings O, Pritchard E, Jones J, House T, et al. Tracking the Emergence of SARS-CoV-2 Alpha Variant in the United Kingdom. N Engl J Med. 2021;385(27):2582–5.

10. Washington NL, Gangavarapu K, Zeller M, Bolze A, Cirulli ET, Schiabor Barrett KM, et al. Emergence and rapid transmission of SARS-CoV-2 B.1.1.7 in the United States. Cell. 2021;184(10):2587–94 e7.

11. Tian F, Tong B, Sun L, Shi S, Zheng B, Wang Z, et al. N501Y mutation of spike protein in SARS-CoV-2 strengthens its binding to receptor ACE2. Elife. 2021;10.

12. Petrone ME, Rothman JE, Breban MI, Ott IM, Russell A, Lasek-Nesselquist E, et al. Combining genomic and epidemiological data to compare the transmissibility of SARS-CoV-2 variants Alpha and Iota. Commun Biol. 2022;5(1):439.

13. Russell A, O’Connor C, Lasek-Nesselquist E, Plitnick J, Kelly JP, Lamson DM, George KS. Spatiotemporal analyses of 2 co-circulating SARS-CoV-2 variants, New York state, USA. Emerging infectious diseases. 2022;28(3), p.650.

14. Turakhia Y, Thornlow B, Hinrichs AS, De Maio N, Gozashti L, Lanfear R, et al. Ultrafast Sample placement on Existing tRees (UShER) enables real-time phylogenetics for the SARS-CoV-2 pandemic. Nat Genet. 2021;53(6):809–16.

15. Katoh K, Asimenos G, Toh H. Multiple alignment of DNA sequences with MAFFT. Methods Mol Biol. 2009;537:39–64.

16. Minh BQ, Schmidt HA, Chernomor O, Schrempf D, Woodhams MD, von Haeseler A, et al. IQ-TREE 2: New Models and Efficient Methods for Phylogenetic Inference in the Genomic Era. Mol Biol Evol. 2020;37(5):1530–4.

17. NYC DOHMH population estimates, modified from US Census Bureau interpolated intercensal population estimates, 2000-2021. Updated September 2022.

18. Suchard MA, Lemey P, Baele G, Ayres DL, Drummond AJ, Rambaut A. Bayesian phylogenetic and phylodynamic data integration using BEAST 1.10. Virus Evol. 2018;4(1):vey016.

19. Hill V, Baele G. Bayesian Estimation of Past Population Dynamics in BEAST 1.10 Using the Skygrid Coalescent Model. Mol Biol Evol. 2019;36(11):2620–8.

20. Rambaut A, Drummond AJ, Xie D, Baele G, Suchard MA. Posterior Summarization in Bayesian Phylogenetics Using Tracer 1.7. Syst Biol. 2018;67(5):901–4.

21. Lemey P, Hong SL, Hill V, Baele G, Poletto C, Colizza V, et al. Accommodating individual travel history and unsampled diversity in Bayesian phylogeographic inference of SARS-CoV-2. Nat Commun. 2020;11(1):5110.

22. Lemey P, Ruktanonchai N, Hong SL, Colizza V, Poletto C, Van den Broeck F, et al. Untangling introductions and persistence in COVID-19 resurgence in Europe. Nature. 2021;595(7869):713–7.

23. Orf GS, Perez LJ, Ciuoderis K, Cardona A, Villegas S, Hernandez-Ortiz JP, et al. The Principles of SARS-CoV-2 Intervariant Competition Are Exemplified in the Pre-Omicron Era of the Colombian Epidemic. Microbiol Spectr. 2023;11(3):e0534622.

24. Vargas-Herrera N, Araujo-Castillo RV, Mestanza O, Galarza M, Rojas-Serrano N, Solari-Zerpa L. SARS-CoV-2 Lambda and Gamma variants competition in Peru, a country with high seroprevalence. Lancet Reg Health Am. 2022;6:100112.

25. Vasylyeva TI, Fang CE, Su M, Havens JL, Parker E, Wang JC, et al. Introduction and Establishment of SARS-CoV-2 Gamma Variant in New York City in Early 2021. J Infect Dis. 2022;226(12):2142–9.

26. Naveca FG, Nascimento V, de Souza VC, Corado AL, Nascimento F, Silva G, et al. COVID-19 in Amazonas, Brazil, was driven by the persistence of endemic lineages and P.1 emergence. Nat Med. 2021;27(7):1230–8.

27. Faria NR, Mellan TA, Whittaker C, Claro IM, Candido DDS, Mishra S, et al. Genomics and epidemiology of the P.1 SARS-CoV-2 lineage in Manaus, Brazil. Science. 2021.

28. Giovanetti M, Fonseca V, Wilkinson E, Tegally H, San EJ, Althaus CL, et al. Replacement of the Gamma by the Delta variant in Brazil: Impact of lineage displacement on the ongoing pandemic. Virus Evol. 2022;8(1):veac024.

29. Dellicour S, Hong SL, Hill V, Dimartino D, Marier C, Zappile P, et al. Variant-specific introduction and dispersal dynamics of SARS-CoV-2 in New York City - from Alpha to Omicron. PLoS Pathog. 2023;19(4):e1011348.

